# Maternal and Neonatal Outcomes in Women with Inflammatory Bowel Disease Following Different Biologic Utilization Trajectories in Pregnancy

**DOI:** 10.64898/2026.01.21.26344506

**Authors:** Elise Van den Broeck, Els De Dycker, Zenobie Annaert, Patricia Geens, Tessy Lambrechts, Elien Loddewijkx, Sarah Brödel, Kristel Van Calsteren, Lore Lannoo, João Pedro Guedelha Sabino, Bram Verstockt, Mette Julsgaard, Marc Ferrante, Michael Ceulemans

**Author notes:** Corresponding author: Michael Ceulemans, PharmD, PhD, Department of Pharmaceutical and Pharmacological Sciences, KU Leuven Campus Gasthuisberg, ON2, Herestraat 49 box 521, 3000 Leuven, Belgium. Joint first authors (Elise Van den Broeck, Els De Dycker). Joint last authors (Marc Ferrante, Michael Ceulemans). **Conflict of interest** The BELpREG pregnancy register, coordinated by the Department of Pharmaceutical and Pharmacological Sciences at KU Leuven (with involvement of M.C. and E.V.D.B.), has received independent research grants and sponsorships for the educational activities from UCB, Almirall, Pfizer, Sanofi, Sandoz and J&J. J.S. has received speaker fees from Pfizer, Abbvie, Ferring, Falk, Takeda, Janssen, Fresenius, and Galapagos; Consultancy fees from Pfizer, Janssen, Ferring, Fresenius, Abbvie, Galapagos, Celltrion, Pharmacosmos, and Pharmanovia; and research support from Galapagos and Viatris. B.V. received research support from AbbVie, Biora Therapeutics, Celltrion, Landos, Pfizer, Sossei Heptares, and Takeda; speaker’s fees from AbbVie, Biogen, Bristol Myers Squibb, Celltrion, Chiesi, Eli Lilly, Falk, Ferring, Galapagos, Johnson and Johnson, MSD, Pfizer, R-Biopharm, Sandoz, Takeda, Tillots Pharma, Truvion, and Viatris; and consultancy fees from AbbVie, Alfasigma, Alimentiv, Applied Strategic, AstraZeneca, Atheneum, BenevolentAI, Biora Therapeutics, Boxer Capital, Bristol Myers Squibb, Eli Lilly, Galapagos, Guidepont, Landos, Merck, Mylan, Nxera, Inotrem, Ipsos, Johnson and Johnson, Pfizer, Progenity, Sandoz, Sanofi, Santa Ana Bio, Sapphire Therapeutics, Sosei Heptares, Takeda, Tillots Pharma, and Viatris; and holds stock options in Vagustim. M.J. received research funding from Takeda; consulting/advisory board fees from Ferring, Takeda, AbbVie, PharmaCosmos, Eli Lilly and Tillots Pharma; and speaker’s fees from Tillotts Pharma, MSD, Ferring, Janssen and Takeda. M.F. reports Research grants from AbbVie, EG Pharma, Celltrion, Janssen, Pfizer, Takeda and Viatris; consultancy fees from AbbVie, AgomAb Therapeutics, Boehringer Ingelheim, Celgene, Celltrion, Eli Lilly, Janssen-Cilag, MRM Health, Merck Sharp and Dohme, Pfizer, Takeda and ThermoFisher; and speakers’ fees from AbbVie, Biogen, Boehringer Ingelheim, Dr Falk Pharma, Ferring, Janssen-Cilag, Merck Sharp and Dohme, Pfizer, Takeda, Truvion Healthcare and Viatris. All above-mentioned companies had no influence on the design, conduct or reporting of this study. The remaining authors have no conflicts of interest to declare. **Data availability statement**: The data supporting the findings of this study are available from the corresponding author upon request.

## Abstract

**Objectives:** The 2025 Global Consensus recommends continuing biologics throughout pregnancy in women with inflammatory bowel disease (IBD). Real-world evidence on biologic treatment patterns and outcomes remains limited. This study compared maternal and neonatal outcomes across different biologic use trajectories during pregnancy.

**Methods:** A retrospective study was performed in pregnant women with IBD, treated and/or delivering at the University Hospitals Leuven, Belgium, between 2017 and 2025. Patients were categorized as continuers, discontinuers, non-users or initiators of biologics during pregnancy

**Results:** Among 255 pregnancies, 103 (40.4%) were continuers, 68 (26.7%) discontinuers, 77 (30.2%) non-users, and 7 (2.7%) initiators. Before conception, 67.1% used biologics. Third-trimester disease activity was most frequent in initiators (42.9%, 3/7) and discontinuers (19.1%, 13/68), followed by non-users (14.3%, 11/77) and continuers (13.6%, 14/103). C-sections occurred more often in non-users (41.3%, 26/63) and discontinuers (39.4%, 26/66) than continuers (31.1%, 23/74). Preterm birth was more common among initiators (14.3%, 1/7) and discontinuers (12.1%, 8/66) than continuers (8.0%, 6/75) and non-users (3.2%, 2/62). Low birthweight occurred most in initiators (14.3%, 1/7), continuers (8.1%, 6/74) and discontinuers (6.1%, 4/66). Small-for-gestational-age infants were most frequent among continuers (14.9%, 11/74) and initiators (14.3%, 1/7) than discontinuers (7.6%, 5/66).

**Conclusions:** Women who discontinued biologics during pregnancy had higher rates of C-sections, preterm birth, and third-trimester disease activity than continuers, supporting continuation of biologics in pregnancy. The higher SGA rates among continuers, however, require further investigation. Initiators showed the poorest outcomes, highlighting the need for adequate disease control before and during pregnancy.

## Introduction

Inflammatory bowel disease (IBD), including Crohn’s disease and ulcerative colitis, affects 7 million people worldwide and the incidence and prevalence are increasing (1–4). These conditions frequently occur in women of childbearing age (5). For those planning pregnancy, remission prior to conception is critical, as active disease confers a fivefold higher risk of flares during pregnancy (6–8) and adverse neonatal outcomes (e.g. spontaneous abortions, preterm delivery, low birthweight and small for gestational age) (5, 9, 10). Biologics are effective for maintaining remission and preventing disease flares and may be required during pregnancy to sustain disease control (11, 12). However, the 2015 European Crohn’s and Colitis Organization (ECCO) guideline recommended discontinuing biologics in the second trimester for patients with sustained remission, reflecting the lack of safety evidence and uncertainty regarding potential teratogenic effects (13). In the last decade, emerging evidence has led to updated recommendations, with the 2022 ECCO guideline supporting continued anti-TNF use in pregnancy and the 2025 Global Consensus guideline supporting the continued use of TNF inhibitors, anti-integrins, and interleukin inhibitors throughout the entire pregnancy (14, 15). Despite these changes, more real-world data on different biologic utilization patterns and their impact on pregnancy and neonatal outcomes are needed (16). This study aimed to investigate maternal and neonatal outcomes following different biologic use trajectories during pregnancy in a tertiary referral center in Belgium.

## Methods

### Study design

This retrospective, observational study reviewed medical records of pregnant women with IBD, who were treated and/or delivered at the tertiary centre of University Hospitals Leuven between 2017 and 2025. Patients were classified into four groups according to their biological use in pregnancy: continuers, discontinuers, non-users and initiators. All patients provided written informed consent for inclusion in the Crohn’s Disease and Ulcerative Colitis Advanced Research (CCARE) database (S53684), authorizing the use of their data for scientific research. The study was approved by the Ethical Committee UZ/KU Leuven (MP022851).

### Data collection

In September 2025, data were retrieved from electronic hospital medical records. Collected information included maternal age at conception, IBD subtype, previous pregnancies and abortions, substance use, folic acid supplementation, clinical documentation of disease activity, preconception IBD treatment, and medication changes during pregnancy. Maternal outcomes included mode of delivery and gestational age at birth (including preterm birth (<37 weeks). Neonatal outcomes comprised birthweight, length, low birthweight (<2500 grams), and small for gestational age (SGA) (calculated using local growth curve data (17)), defining SGA as <10^th^ birth weight percentile according to gestational age at delivery and infant sex.

Preconception disease activity was identified when noted in the medical record. Disease activity during pregnancy was assessed using a composite endpoint, defined as the notification of active disease in the medical record, or initiation or alteration of IBD-related therapy, or the delivery prompted by disease activity. Active disease was not assumed when medication changes were solely related to the discontinuation of clinical trial medication.

To ensure consistency in group classification, biologic continuation was defined as uninterrupted use throughout pregnancy, regardless of changes in non-biologic IBD medications. Switching between biologic agents within the same therapeutic class did not alter group assignment. Changes in non-biologic medications were disregarded for allocation purposes. Discontinuers were defined as patients who discontinued biologics at any point during pregnancy, irrespective of any reinitiation later in pregnancy.

### Data analysis

Descriptive statistics were applied and expressed as absolute numbers and percentages. Missing data were excluded so that percentages reflected available data only. For women who discontinued or initiated biologic therapy, the timing of these changes was determined. The final week of the preceding trimester was used in the analysis when it was documented in the records to have occurred before the start of a specific trimester. Trimesters were defined based on gestational age starting from the first day of the last menstrual period: week 0-12 (first trimester), week 13-26 (second trimester), and week 27 until delivery (third trimester).

Maternal and neonatal outcomes were compared across the four biologics trajectory groups. Moreover, maternal and neonatal outcomes were also compared according to the timing of delivery, i.e. before or after the implementation of the 2022 ECCO guideline in our center. Therefore, group 1 included patients who delivered before January 1^st^, 2023, while group 2 comprised patients who delivered from January 1^st^, 2023, onwards. The cut-off was chosen more arbitrarily but taking into account the publication of the 2022 ECCO guideline and the anticipated implementation in our center. In both analyses, continuous variables for continuers, discontinuers and non-users were analyzed using ANOVA when the assumptions of normality and homogeneity of variances were met. Otherwise, the Kruskal-Wallis test was performed. Categorical variables were analyzed using Chi-square tests, or Fisher’s Exact test when expected cell counts were below five. Statistical significance was defined as a p-value <0.05. Outcomes for initiators were summarized descriptively due to the limited sample size of that group. All statistical analyses were performed with R (version 2025.08.0+345).

## Results

### Characteristics of participants across the different group

A total of 255 pregnancies in 176 women were analyzed: 103 (40.4%) were continuers, 68 (26.7%) discontinuers, 77 (30.2%) non-users, and 7 (2.7%) initiators, as summarized in Table 1. In 20 of these 255 pregnancies (7.8%), medication changes occurred without affecting group allocation (16 due to initiation of conventional IBD therapy, 1 due to discontinuation of conventional therapy, and 3 due to switches between biologics from the same therapeutic class). Biologics were typically discontinued or initiated at 22 weeks of gestation.

**Table 1.** Group Allocation Based on Biologic Treatment Trajectory During Pregnancy the distribution of 255 pregnant women across four categories based on their use of biologics during pregnancy. Discontinuation or initiation before a specific trimester was interpreted as occurring in the last week of the preceding trimester. Trimesters were defined as follows: week 0-12 (first trimester), week 13-26 (second trimester), and week 27 until delivery (third trimester).

| Table 1. Group Allocation Based on Biologic Treatment Trajectory During Pregnancy |  |  |
| --- | --- | --- |
| Group | Description | Frequency (%) |
| Group 1: Continuers | Biological therapy continued until delivery | 103/255 (40.4) |
| Group 2:<br>Discontinuers | Biological therapy discontinued during pregnancy | 68/255 (26.7) |
|  | First trimester | 1/68 (1.5) |
|  | Second trimester | 57/68 (83.8) |
|  | Third trimester | 3/68 (4.4) |
|  | Timing not specified | 7/68 (10.3) |
| Group 3: Non-users | No biological therapy used during the entire pregnancy | 77/255 (30.2) |
| Group 4: Initiators | Biologics initiated during pregnancy | 7/255 (2.7) |
|  | First trimester | 1/7 (14.3) |
|  | Second trimester | 1/7 (17.3) |
|  | Third trimester | 2/7 (28.6) |
|  | Timing not specified | 3/7 (42.9) |
| Mean (±SD) gestational week of biologic therapy discontinuation (group 2) (n=59) | | 22.1 (±2.4) |
| Mean (±SD) gestational week of biologic therapy initiation (group 4) (n=4) | | 22.5 (±3.0) |

Mean maternal age at conception and IBD subtype distribution were similar across groups. Disease activity during pregnancy was highest among initiators (100.0%, 7/7), followed by discontinuers (27.9%, 19/68), continuers (22.3%, 23/103), and non-users (19.5%, 15/77), and was generally most commonly reported in the third trimester (i.e., initiators (42.9%, 3/7), discontinuers (19.1%, 13/68), non-users (14.3%, 11/77), and continuers (13.6%, 14/103)). Before conception, 67.1% used biologics, predominantly infliximab, vedolizumab, adalimumab, and ustekinumab (see Table 2).

**Table 2.** Baseline and treatment characteristics by exposure group presents the baseline characteristics of 255 pregnancies in 176 women with inflammatory bowel disease. Conventional therapy consisted of aminosalicylates, corticosteroids and immunosuppressives. Biologics included TNF-alpha inhibitors, anti-integrins and interleukin inhibitors, and small molecules included JAK-inhibitors and sphingosine-1-phosphate receptor modulators. Percentages are calculated based on the number of participants with available data for each parameter. Disease activity during pregnancy was defined as a composite variable based on the notification of active disease in the medical record, or initiation or alteration of IBD-related therapy, or the delivery prompted by disease activity. For disease activity, the combined trimester counts may exceed the overall number of women with disease activity during pregnancy, as a single pregnancy could have activity in multiple trimesters. Women in group 1 continued biologics throughout the entire pregnancy. Group 2 discontinued biologics at some point during pregnancy. Group 3 did not use any biologics during pregnancy, while group 4 initiated biologics during pregnancy.

| Maternal information |  |  |  |  |  |
| --- | --- | --- | --- | --- | --- |
|  | **Group 1: Continuers**<br>(n=103) | **Group 2: Discontinuers**<br>(n=68) | **Group 3: Non-Users**<br>(n=77) | **Group 4: Initiators**<br>(n=7) | **Total**<br>(n=255) |
| Maternal age at conception (mean±SD) (years) | 30.9±4.3 | 30.6±4.4 | 30.7±4.3 | 29.7±4.8 | 30.6±4.3 |
| IBD subtype (%) |  |  |  |  |  |
| Crohn's disease | 66/103 (64.1) | 47/68 (69.1) | 44/77 (57.1) | 4/7 (57.1) | 161/255 (63.1) |
| Ulcerative colitis | 37/103 (35.9) | 21/68 (30.9) | 32/77 (41.6) | 3/7 (42.9) | 93/255 (36.5) |
| IBD type unclassified | 0/103 (0.0) | 0/68 (0.0) | 1/77 (1.3) | 0/7 (0.0) | 1/255 (0.4) |
| Substance use during pregnancy (%) |  |  |  |  |  |
| Alcohol | 3/63 (4.8) | 0/42 (0.0) | 2/46 (4.3) | 0/7 (0.0) | 5/155 (3.2) |
| Tobacco | 11/98 (11.2) | 7/65 (10.8) | 4/75 (5.3) | 2/7 (28.6) | 24/246 (9.8) |
| Folic acid use during pregnancy (%) | 84/85 (98.8) | 59/59 (100.0) | 63/63 (100.0) | 6/6 (100.0) | 213/214 (99.5) |
| Pregnancy history (%) |  |  |  |  |  |
| ≥1 prior pregnancy | 45/99 (45.5) | 46/67 (68.7) | 44/77 (57.1) | 5/7 (71.4) | 140/250 (56.0) |
| ≥1 abortions | 22/45 (48.9) | 25/46 (54.3) | 18/44 (40.9) | 3/5 (60.0) | 68/140 (48.6) |
| Active disease (%) |  |  |  |  |  |
| Preconception | 12/103 (11.7) | 5/68 (7.4) | 8/62 (12.9) | 0/2 (0.0) | 25/235 (10.6) |
| During pregnancy | 23/103 (22.3) | 19/68 (27.9) | 15/77 (19.5) | 7/7 (100.0) | 64/255 (25.1) |
| First trimester | 8/103 (7.8) | 6/68 (8.8) | 4/77 (5.2) | 4/7 (57.1) | 22/255 (8.6) |
| Second trimester | 3/103 (2.9) | 3/68 (4.4) | 5/77 (6.5) | 6/7 (85.7) | 17/255 (6.7) |
| Third trimester | 14/103 (13.6) | 13/68 (19.1) | 11/77 (14.3) | 3/7 (42.9) | 41/255 (16.1) |
| Not specified | 1/103 (1.0) | 0/68 (0.0) | 0/77 (0.0) | 0/7 (0.0) | 1/255 (0.4) |
| Preconception IBD-related treatment |  |  |  |  |  |
| Conventional therapy (%) | 15/103 (14.6) | 11/68 (16.4) | 38/77 (48.7) | 1/7 (14.3) | 65/255 (25.5) |
| Biologics (%) | 103/103 (100) | 68/68 (100) | 0/77 (0.0) | 0/7 (0.0) | 171/255 (67.1) |
| Infliximab | 35/103 (34.0) | 17/68 (25.0) | 0/77 (0.0) | 0/7 (0.0) | 52/255 (20.4) |
| Vedolizumab | 24/103 (23.3) | 20/68 (29.4) | 0/77 (0.0) | 0/7 (0.0) | 44/255 (17.3) |
| Adalimumab | 21/103 (20.4) | 15/68 (22.1) | 0/77 (0.0) | 0/7 (0.0) | 36/255 (14.1) |
| Ustekinumab | 16/103 (15.5) | 12/68 (17.6) | 0/77 (0.0) | 0/7 (0.0) | 28/255 (11.0) |
| Risankizumab | 5/103 (4.9) | 1/68 (1.5) | 0/77 (0.0) | 0/7 (0.0) | 6/255 (2.4) |
| Study medication | 1/103 (1.0) | 2/68 (2.9) | 0/77 (0.0) | 0/7 (0.0) | 3/255 (1.2) |
| Golimumab | 1/103 (1.0) | 1/68 (1.5) | 0/77 (0.0) | 0/7 (0.0) | 2/255 (0.8) |
| Small molecules | 0/103 (0.0) | 0/68 (0.0) | 0/77 (0.0) | 0/7 (0.0) | 0/255 (0.0) |

The distribution of biological classes used during pregnancy remained similar to the preconception period. One woman switched in pregnancy to mirikizumab following discontinuation of study medication, while another woman switched from adalimumab to infliximab. Among initiators, four women (57.1%, 4/7) started infliximab at a mean gestational age of 26.7 weeks, one (14.3%, 1/7) initiated adalimumab at 10 weeks’ gestation, and vedolizumab and ustekinumab were each initiated by one woman (14.3%, 1/7).

Active disease during pregnancy was reported in 64 (25.1%) of all pregnancies. A total of 37 (14.5%) pregnancies involved medication changes due to increased disease activity during pregnancy. Of these, 28 women initiated conventional therapy during pregnancy (nine were continuers, eight discontinuers, ten were non-users and none were initiators), including corticosteroids (n=15), mesalamine (n=13), azathioprine (n=2), and antibiotics (n=1) - two patients initiated both corticosteroids and mesalamine and one initiated azathioprine and corticosteroids during pregnancy. Of the other nine women, five received biologics (four initiators and one discontinuer who eventually restarted ustekinumab due to disease activity), and four received a combination of conventional IBD therapy and biologics (three initiators and one discontinuer who restarted adalimumab due to disease activity).

### Maternal and neonatal outcomes across different biologic use trajectories

The mean (SD) gestational age at delivery was 38.9 (2.0) weeks, with no difference between groups (p=0.26) (see Table 3). C-section rates were highest in initiators (42.9%, 3/7), non-users (41.3%, 26/63) and discontinuers (39.4%, 26/66), compared to continuers (31.1%, 23/74), with an observed difference across groups (p=0.005). Preterm birth was most common in initiators (14.3%, 1/7) and discontinuers (12.1%, 8/66), followed by continuers (8.0%, 6/75) and non-users (3.2%, 2/62), with a borderline significant difference across groups (p=0.10). Non-users showed a higher birth weight (3366g) compared to initiators (2999g), continuers (3206g) and discontinuers (3239g), also with a borderline significant difference across groups (p=0.09). Low birthweight occurred most often in initiators (14.3%, 1/7), compared to continuers (8.1%, 6/74), discontinuers (6.1%, 4/66) and non-users (1.6%, 1/62) (p=0.21). SGA was more common in continuers (14.9%, 11/74) and initiators (14.3%, 1/7), than in discontinuers (7.6%, 5/66) and non-users (6.3%, 4/63) (p=0.23). Of the 21 women with SGA, 7 had disease activity during pregnancy (33.3%). Among continuers, only 2 of the 11 women with SGA (18.2%) had active disease, whereas this was the case for 3 of the 5 women (60.0%) who discontinued their biologic.

**Table 3.** Mother-Child Outcomes subdivided per group summarizes pregnancy and neonatal outcomes of 255 pregnancies in 176 IBD patients. Percentages are calculated based on the number of participants with available data for each parameter. P-values are calculated for the comparison of group 1 (continuers), group 2 (discontinuers) and group 3 (non-users). P-values for continuous variables are calculated using ANOVA. For gestational age at delivery, a Kruskal-Wallis test has been used. Categorical variables are analyzed using Fisher’s Exact test, except for method of delivery, which has been assessed with a Chi-square test. Women in group 1 continued biologics throughout the entire pregnancy. Group 2 discontinued biologics at some point during pregnancy. Group 3 did not use any biologics during pregnancy, while group 4 initiated biologics during pregnancy.

| Table 3. Mother-Child Outcomes subdivided per group |  |  |  |  |  |  |
| --- | --- | --- | --- | --- | --- | --- |
|  | **Group 1:**<br>**Continuers**<br>**(n=103)** | **Group 2:**<br>**Discontinuers**<br>**(n=68)** | **Group 3:**<br>**Non-users**<br>**(n=77)** | **P-value** | **Group 4:**<br>**Initiators**<br>**(n=7)** | **Total**<br>**(n=255)** |
| Maternal outcomes |  |  |  |  |  |  |
| Method of delivery (%) |  |  |  |  |  |  |
| Vaginal | 51/74 (68.9) | 40/66 (60.6) | 37/63 (58.7) | 0.005 | 4/7 (57.1) | 132/210 (62.9) |
| Caesarean section | 23/74 (31.1) | 26/66 (39.4) | 26/63 (41.3) |  | 3/7 (42.9) | 78/210 (37.1) |
| Primary section | 14/23 (60.9) | 17/26 (65.4) | 15/26 (57.7) |  | 2/3 (66.7) | 48/78 (61.5) |
| Secondary section | 9/23 (39.1) | 9/26 (34.6) | 8/26 (30.8) |  | 1/3 (33.3) | 27/78 (34.6) |
| Unknown | 0/23 (0.0) | 0/26 (0.0) | 3/26 (11.5) |  | 0/3 (0.0) | 3/78 (3.8) |
| Mean (±SD) gestational age at delivery (weeks) | 39.0±2.0 | 38.6±2.1 | 39.1±2.0 | 0.26 | 39.4±1.8 | 38.9±2.0 |
| Neonatal outcomes |  |  |  |  |  |  |
|  | n=77 | n=66 | n=63 |  | n=7 | n=213 |
| Preterm birth (<37w gestation) (%) | 6/75 (8.0) | 8/66 (12.1) | 2/62 (3.2) | 0.10 | 1/7 (14.3) | 17/210 (8.1) |
| Length (mean±sd) (centimeter) | 49.7±2.6 | 48.8±3.7 | 49.9±2.1 | 0.47 | 48.4±2.4 | 49.1±2.1 |
| Birthweight (mean±sd) (grams) | 3206.3±480.0 | 3239.3±480.9 | 3366.2±482.9 | 0.09 | 2998.6±469.5 | 3257.2±459.1 |
| Low birthweight (<2500g) (%) | 6/74 (8.1) | 4/66 (6.1) | 1/62 (1.6) | 0.21 | 1/7 (14.3) | 12/209 (5.7) |
| Small for gestational age (%) | 11/74 (14.9) | 5/66 (7.6) | 4/62 (6.5) | 0.23 | 1/7 (14.3) | 21/209 (10.0) |

### Maternal and neonatal outcomes according to the timing of delivery

The additional analysis showed substantial differences in biologic use patterns between the two study periods (see Table 4). In group 1 (n=124) (i.e., all deliveries before January 1^st^, 2023), only 23 women (18.5%) continued biologic use, while 61 (49.2%) discontinued biologics in pregnancy. 37 (29.8%) were non-users and 3 (2.4%) initiated biologics. In contrast, group 2 (n=87) (i.e., all deliveries from January 1^st^, 2023, onwards), consisted of 52 (59.8%) continuers and only 5 (5.7%) discontinuers. 26 (29.9%) were non-users and 4 (4.6%) initiators.

**Table 4.**
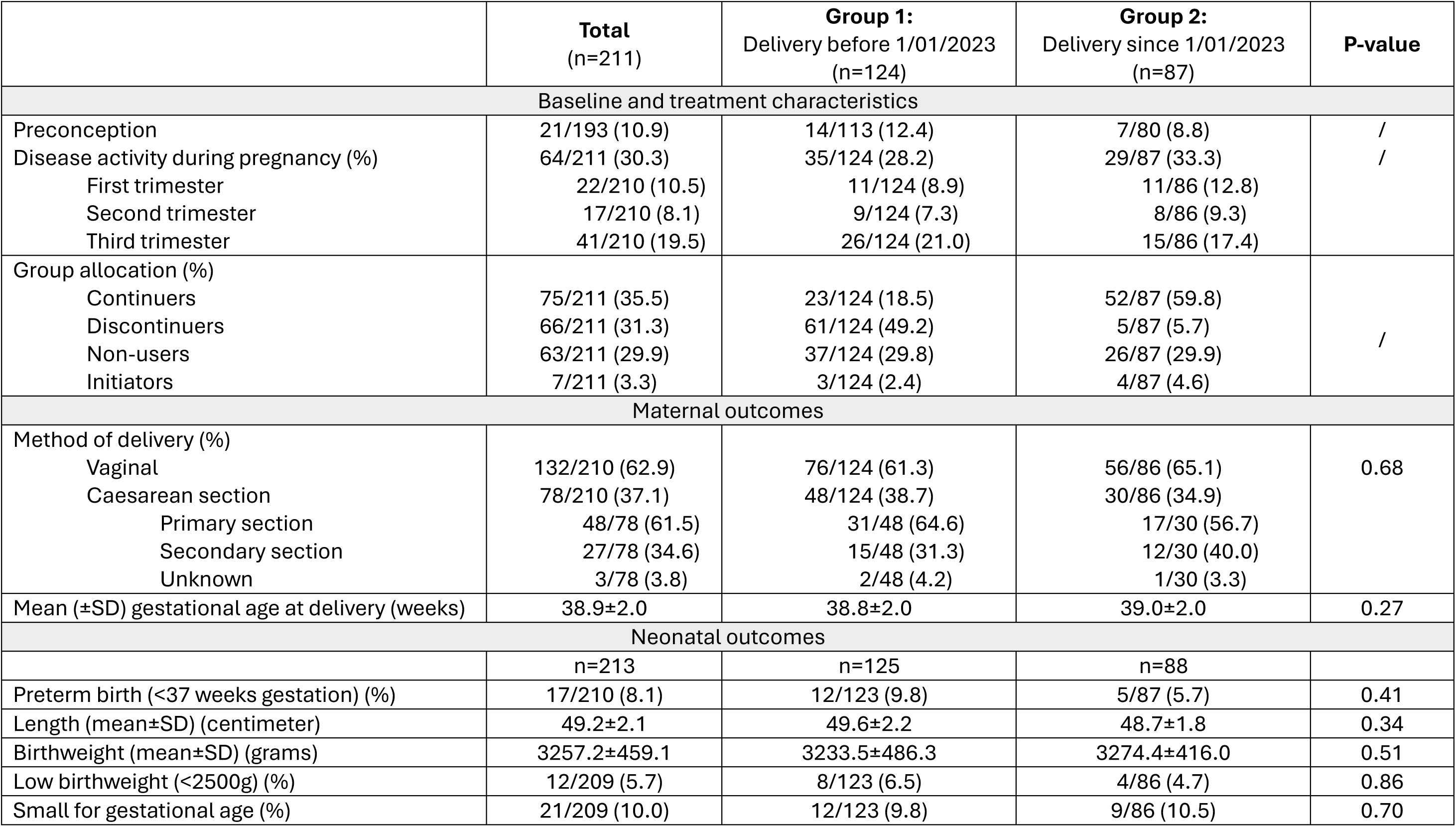
Mother-Child Outcomes according to the timing of the delivery presents pregnancy and neonatal outcomes of pregnancies who ended with a live birth before or from January 1^st^, 2023, onwards. Percentages are calculated based on the number of participants with available data for each parameter. P-values for continuous variables are calculated using ANOVA. For gestational age at delivery, a Kruskal-Wallis test has been used. Categorical variables are analyzed using Chi-square tests, except for low birthweight, which has been assessed with a Fisher’s Exact test.

Overall, third-trimester disease activity was somewhat lower in group 2 (17.4%, 15/86) compared to group 1 (21.0%, 26/124). C-section rates were lower in group 2 (34.9%, 30/86) than in group 1 (38.7%, 48/124) (p=0.68). Preterm birth also occurred less often in group 2 (5.7%, 5/87) compared to group 1 (9.8%, 12/123) (p=0.41). Similarly, low birthweight was less common in group 2 (4.7%, 4/86) than in group 1 (7.3%, 9/124) (p=0.86). SGA infants were, however, slightly more frequent in group 2 (10.5%, 9/86) than in group 1 (9.8%, 12/123) (p=0.70).

## Discussion

### Main findings

This study aimed to assess maternal and neonatal outcomes following different biologic use trajectories during pregnancy. In this IBD cohort from a tertiary hospital in Belgium, 10.6% of women were not in remission at the time of conception and one in four had disease activity during pregnancy. Notably, all women who initiated biologics during pregnancy did so in the context of active disease. Among women who discontinued biologics, up to 30% experienced disease activity, most often in the third trimester (19.1%), whereas continuing biologics throughout pregnancy showed lower third-trimester disease activity (13.6%). This cessation pattern aligns with the 2015 European guideline advising discontinuation of biologics in the second trimester (13) but is not in line with the most recent consensus guideline (6) and may carry risks for active disease later in pregnancy and adverse mother-child outcomes.

In our cohort, C-section was more common in discontinuers compared with continuers, although obstetric factors unrelated to IBD may have contributed. Other outcomes did not statistically differ between both groups. However, preterm birth occurred less often in continuers compared to discontinuers and initiators, both groups with the highest rates of disease activity in the third trimester. This finding aligns well with previous evidence showing higher preterm birth rates in women with active disease (10, 11, 14, 18–21). Surprisingly, SGA infants were more common among continuers. However, continuers with an SGA pregnancy were much less likely to have active disease during pregnancy compared with discontinuers with an SGA pregnancy. Further studies are needed to determine what explains this higher proportion of SGA infants in continuers, such as potential unmeasured disease severity or the extent of the disease in this group. As two third of our cohort used biologics before conception, this tertiary-center population likely reflects the inclusion of more severe IBD patients. Previous studies have shown increased disease activity during pregnancy and postpartum when TNF inhibitors were discontinued (6, 7, 22), while continuation during pregnancy has not been linked to adverse outcomes such as preterm birth or SGA (6, 18, 23). Finally, all initiators had active disease during pregnancy and exhibited the highest rates of preterm birth and C-sections, emphasizing the importance of adequate disease control throughout pregnancy.

With respect to short term maternal and infant outcomes, our findings support the continuation of biologics throughout pregnancy, as maintaining therapy reduces the risk of active disease and related adverse outcomes. However, it is important to acknowledge that the currently available safety evidence in pregnancy varies across biologics. While adalimumab, infliximab, ustekinumab and vedolizumab have been more studied, safety data for newer agents such as mirikizumab and risankizumab are very limited. The continuation of biologics has now been recommended by ECCO and PIANO (6, 14), as the perceived benefits of biologic continuation outweigh theoretical risks, leading to greater acceptance of ongoing treatment during pregnancy (10, 11, 20, 24). This trend is reflected in recent real-word data from the United States, showing increased biologic use in pregnancy over the last decade (25). Similarly, in our tertiary center, the continuation of biologics increased markedly after the in-house implementation of the 2022 ECCO guideline, with most women from 2023 onward maintaining biologics throughout pregnancy.

Questions about medication safety during pregnancy, combined with the potential risk of active disease at the time of conception and during gestation, highlight the importance of preconception counseling to achieve remission and address medication-related concerns (6, 14, 26). In our center, a dedicated care pathway of preconception and pregnancy counseling of IBD patients has recently been established (26), while its effectiveness on mother-infant outcomes will be further assessed.

### Strengths and limitations

This study offers several strengths. First, the study uses real-world data providing detailed insights into the actual utilization of biologics in pregnant women with IBD in daily clinical practice. By categorizing patients into four distinct groups, the analysis captures different treatment patterns among individuals alongside their maternal and neonatal outcomes.

Second, the inclusion of data collected over multiple years enables the description of a cohort of >250 pregnancies. Third, disease activity was assessed using a composite endpoint incorporating multiple parameters, enabling the most accurate possible estimation throughout pregnancy.

However, some limitations should be acknowledged. First, as the study relied on medical records, the analysis was dependent on the completeness and quality of the documented information and data availability for specific variables. For example, no information was available on patient’s historical use of biologics or on disease severity in the years prior to the index pregnancy, and objective parameters of disease activity, such as fecal calprotectin levels, endoscopic assessment or intestinal ultrasound, were generally lacking. Second, the descriptive design precludes establishing causal relationships between biologic exposure and mother-infant outcomes. Third, the sample size may have reduced the power to detect a significant effect between continuers and discontinuers. The small number of women initiating biologics in pregnancy limited the ability to draw conclusions for the subgroup of initiators. Fourth, confounding by indication remains a challenge, as women with severe disease are more likely to receive advanced (biologic) therapies, making it difficult to distinguish the effects of disease activity from medication exposure. Fifth, the assessment of SGA relied on older reference data from our country (published in 2009). Finally, this study did not include (much) data for all biologics, in particular not for newer agents. The findings of our study therefore primarily pertain to TNF inhibitors, ustekinumab, and vedolizumab. Consequently, published safety data in pregnancy still remain insufficient for some biologics and more real-world data urgently need to be collected through dedicated pregnancy registries (27).

## Conclusion

This cohort from a tertiary referral IBD clinic found that women who discontinued biologics during pregnancy had higher rates of C-sections, preterm birth, and third trimester disease activity compared to continuers, supporting the potential benefits of maintaining biologic therapy during pregnancy. However, the higher rate of SGA infants among continuers warrants further investigation. All initiators of biologics during pregnancy presented with active disease and experienced the highest rates of preterm birth and C-sections, underscoring the importance of disease control during pregnancy.

## Data Availability

The data supporting the findings of this study are available from the corresponding author upon request.

## Acknowledgments

We sincerely thank the Belgian IBD patient organizations, CCV and Crohn-RCUH, for their valuable support. We also gratefully acknowledge Sien Lenie for her contributions to our research initiatives on pregnancy and inflammatory bowel disease. In addition, we thank biostatistician Jose Alejandro Rozo Posada for providing valuable statistical input for our analyses.

